# Colonization Amplification despite Limited In-Hospital Transmission: Modeling the Epidemiological Paradox of *C. difficile* and the Impact of Control Strategies in Healthcare Settings

**DOI:** 10.1101/2025.10.09.25337452

**Authors:** Daniel De-la-Rosa-Martinez, Travis C. Porco, Ashley Hazel, Xinran Liu, Karim Khader, Seth Blumberg

**Author notes:** **Corresponding author: Daniel De-la-Rosa-Martinez, MD, PhD**, Francis I. Proctor Foundation, University of California, San Francisco, **Address**: 490 Illinois St, San Francisco, CA 94158, **Institutional email**, **Personal email**.

## Abstract

**Background:** Although healthcare-associated transmission of *C. difficile* is a recognized public health concern, healthcare-onset infections (CDI) remain comparable in number to community-onset cases. This paradox likely reflects the underappreciated interplay between these settings. We aimed to quantify in-hospital transmission and the hospital’s contribution to community colonization by estimating the intrinsic reproduction number (R_i_) and introducing the colonization amplification index (A_i_), defined as the ratio of colonized patients at discharge to those at admission. Given the potential contribution of external cases, we also evaluated interventions targeting asymptomatic carriers at admission to reduce disease burden.

**Methods and Findings:** We developed a compartmental model informed by data from UCSF Medical Center to capture *C. difficile* transmission dynamics among symptomatic and asymptomatic patients. Across simulations, the median R_i_ was 0.58 (Q1–Q3: 0.50–0.65), consistently indicating limited sustained in-hospital transmission (R_i_<1). In contrast, Ai was 1.9 (Q1–Q3: 1.7–2.1), suggesting substantial amplification of colonization during hospital stay. Sensitivity analyses showed that estimates were mainly influenced by discharge rates, antibiotic exposure, transmission rate, and case classification thresholds. Redefining hospital-onset CDI using thresholds from ≥1 to ≥5 days post-admission increased A_i_ and R_i_ by 13% and 17%, and reduced them by 5% and 9%, respectively. Interventions targeting asymptomatic carriers through contact precautions and/or prophylactic treatment reduced both A_i_ and CDI incidence, with combined interventions yielding the greatest reductions, followed by contact precautions alone.

**Conclusions:** Our findings indicate that in-hospital transmission of *C. difficile* is limited (R_i_<1) and likely sustained by continuous importation of cases from the community. Nevertheless, hospitalization amplifies colonization (A_i_>1), further contributing to community transmission. These results underscore the importance of interventions addressing asymptomatic carriers, a currently overlooked source of spread. Our study highlights the need to broaden metrics beyond R_i_ to capture hospitals’ contribution to the *C. difficile* burden. Future infection control strategies should address colonization dynamics at admission and potentially at discharge to mitigate transmission and reduce the overall burden of *C. difficile*.

## INTRODUCTION

Mathematical models of *C. difficile* transmission have traditionally focused on the within-hospital dynamics of healthcare-associated infection and the impact of control interventions [1,2]. Relatively less attention has been paid to the interplay between community- and hospital-based transmission dynamics, despite growing evidence that asymptomatic carriers and community-imported cases play a critical role in sustaining transmission [1,3–6]. Colonized patients discharged from healthcare settings can potentially introduce the pathogen into the community, while individuals colonized or infected outside the hospital can reintroduce *C. difficile* into healthcare facilities, creating a feedback loop that may contribute to infection persistence [7]. This study explores this relationship by developing a compartmental model informed by clinical data from a university-affiliated medical center.

Traditional transmission models often rely on the reproduction number, defined as the average number of secondary cases generated per index case, to quantify in-hospital transmission. However, this metric typically assumes a closed population and does not account for community importation of cases or transmission that occurs after discharge, thus limiting its ability to reflect the role of healthcare institutions in sustaining broader community transmission [8–11]. To address this, we introduce the amplification index (A_i_) as the ratio of colonized patients at discharge to those at admission. This index serves as a complementary metric to the reproduction number, capturing both in-hospital transmission and the hospital’s contribution to community spread.

A second challenge of quantifying transmission relates to how symptomatic infections are classified as community-onset (CO-CDI) and healthcare-associated *C. difficile* infection (HCA-CDI). Traditionally, this classification relies on heuristic, time-based definitions that assign the infection’s origin based on the timing of symptom onset relative to admission [12]. The higher the proportion classified as CO-CDI, the lower the estimate of health-care-associated transmission. Here, we assess how sensitive transmission estimates are to these thresholds.

Our observation that the community-healthcare interface heavily influences the burden of HCA-CDI motivates an evaluation of control strategies focused on controlling transmission due to the importation of asymptomatic carriers of *C. difficile*. We specifically assess the impact of screening asymptomatic carriers on admission so that targeted contact precautions and/or prophylactic decolonization can be deployed. Our focus on interventions that control asymptomatic transmission addresses an important research gap, given that current infection prevention strategies focus almost exclusively on symptomatic cases, despite substantial evidence that asymptomatic individuals can act as reservoirs for onward transmission in healthcare facilities and the community-at-large [12–16].

## METHODS

While the term *infected* can be used in the literature to denote the presence of bacteria in the body regardless of clinical symptoms, in this work, we use the term specifically to refer to individuals who exhibit symptomatic infection and test positive for *C. difficile* by PCR. While this definition may include some patients with diarrhea unrelated to *C. difficile*, it prioritizes more robust evaluation of transmission.

### Model description

We model *C. difficile* hospital transmission using a deterministic compartmental framework that includes eight compartments: 1) Non-susceptible (R) patients whose gut microbiota has not been disrupted by antibiotics. 2) Susceptible (S) individuals with disrupted microbiota due to antibiotic exposure, who are at risk of *C. difficile* colonization. 3) Short-term asymptomatic carriers (E) or patients colonized with toxigenic *C. difficile*, who may progress to symptomatic infection during their hospital stay. 4) Long-term asymptomatic carriers (C) or patients with persistent colonization who will remain asymptomatic during their hospital stay. 5) Infected (I) individuals or patients with symptomatic *C. difficile* infection, confirmed by PCR. 6) Diagnosed infected (A) individuals who are on contact precautions and antibiotic treatment. Additionally, to explore the potential benefits of contact precautions on asymptomatic carriers, we include two compartments: (7) Diagnosed and isolated asymptomatic carriers type E (K_1_) and (8) type C (K_2_). As part of the intervention, these carriers may be treated and either move to the susceptible compartment or remain colonized if treatment is unsuccessful or not administered. Movement rates between compartments are illustrated in Fig 1.

**Fig 1.**
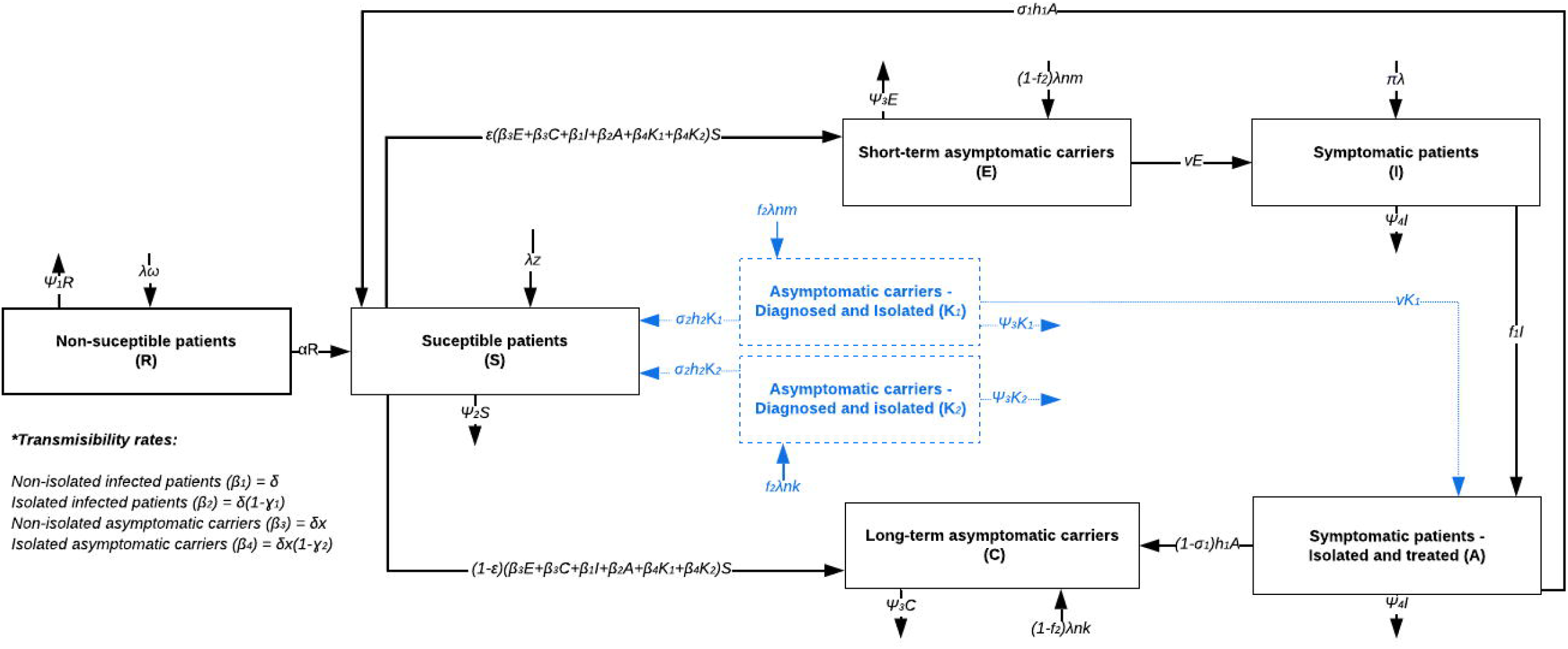
Compartmental model workflow of disease dynamics. Compartments and arrows in black depict the infection dynamics under standard conditions or the baseline transmission model. In contrast, blue-dotted compartments (K_1-2_) and arrows represent hypothetical compartments based on control interventions for asymptomatic carriers, including contact precautions and decolonization.

### Rates, assumptions, and parameters

#### 1.1 Movement from non-susceptible (R) to susceptible (S) compartment

Antibiotic use affects the transition from the non-susceptible to the susceptible population. These agents disrupt the gut microbiome and increase vulnerability to *C. difficile*. Our model uses the α parameter to capture exposure to high-risk antibiotic classes during hospitalization, including quinolones, cephalosporins, sulfonamides, penicillins, macrolides, aminoglycosides, and lincosamides. Therefore, the transition from the R → S compartment is denoted as αR.

#### 1.2 Force of infection of colonized and infected individuals

Susceptible patients can acquire infection from both symptomatic and asymptomatic carriers. The transmission rate of infected individuals is denoted as *δ*. We assume that this rate is reduced by contact precautions, which are typically imperfect due to variable compliance and intrinsic limitations. We denote the overall effectiveness of contact precautions for infected individuals as ɣ_1_. Therefore, the transmissibility rate of infected individuals under contact precautions (compartment A) is calculated as the product of the transmission rate of the infected population (*δ*) and one minus contact precaution effectiveness (ɣ_1_). We use the terms β_1_ and β_2_, where:

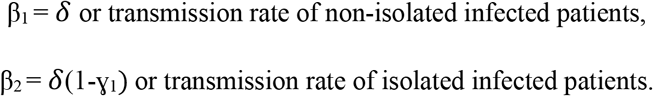

Based on previous evidence, we assume that asymptomatic patients can transmit the infection at a lower rate than patients manifesting active symptoms [14]. In our model, this reduction in transmissibility is indicated by the parameter *x*, which represents the relative transmissibility of the asymptomatic carrier population. Similar to the infected population, the transmissibility of the asymptomatic carrier population can be modified by contact precautions. Therefore, the transmission rate for non-isolated asymptomatic carriers is defined as *δx*, while the asymptomatic carrier population on contact precaution has a transmission rate denoted as *δ*x(1-ɣ_2_), which depends on the effectiveness of contact precaution on asymptomatic carriers, represented as ɣ_2_. We define:

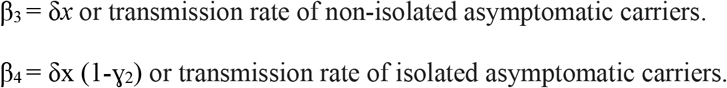

#### 1.3 Movement from susceptible (S) to colonized (E or C) compartments

Susceptible patients can transition into one of two distinct types of asymptomatic carriers: those in the incubation period who may develop symptomatic disease (E compartment) during hospitalization and those who remain asymptomatic throughout their hospital stay (C compartment). Although the factors influencing this progression rate are not fully understood, they may be related to immunological response, microbiome diversity, or *C. difficile* virulence characteristics [5]. Accordingly, the relative movement into each compartment is modeled by the parameter ε, representing the proportion of newly colonized patients who may develop symptomatic disease and enter compartment E.

#### 1.4 Movement from the pre-symptomatic (E) to the infected compartment (I) and the treated/isolated compartment (A)

We used the parameter *v* to represent the rate of symptom development, calculated as the inverse of the days from bacterium acquisition to onset of symptoms. Thus, the transition rate from the E → I compartment is represented as *v*E. Once patients develop symptomatic disease, they are diagnosed at a rate of f_1_ and move from the I → A compartment.

Patients complete treatment at a rate of h_1_, the inverse duration of antibiotics recommended for infection v therapy. Following treatment, a proportion σ_1_ of patients fully clear the bacteria and transition to the S compartment, reflecting treatment effectiveness. In contrast, the remaining proportion, 1 – σ_1_, does not clear the infection and remains colonized, moving to the C compartment. Therefore, σ_1_h_1_ denotes the transition rate from the A→S, and (1-σ_1_)h_1_ represents the movement from the A → C compartment. Although some individuals who do not fully clear the bacteria may experience relapse of infection, such episodes typically occur after discharge and were not included in this model.

#### 1.5 Admission rate and community sources of non-susceptible, susceptible, asymptomatic, and infected patients

The overall admission rate is represented by λ, derived from clinical data as the average daily number of admissions at the institution during the study period. The proportion of patients admitted from the community with symptomatic infection or asymptomatic colonization is denoted as π and n, respectively. A fraction m of asymptomatic carriers is assumed to be at risk for symptomatic infection during hospitalization and enter the E compartment. The remaining asymptomatic carriers, 1-m (denoted as k), stay asymptomatic and enter the C compartment. The proportion of patients, z, who received antibiotics before admission but did not carry *C. difficile* enter the S compartment. The proportion (1 – (n + π + z)), denoted as ω, enters the R compartment.

#### 1.6 Simulated treatment and isolation of asymptomatic carriers

We modeled active screening and isolation of asymptomatic carriers at admission through compartments K_1_ and K_2_. The parameter f_2_ defines the proportion of carriers identified upon admission. As a result, f_2_λnm and f_2_λnk represent the rates at which patients initially assigned to E and C are instead moved to K_1_ and K_2_, respectively. The remaining carriers, those not identified, enter E and C at rates (1 – f_2_)λnm and (1 – f_2_)λnk.

Patients in K_1_ and K_2_ are placed under contact precautions until discharge. If treatment is initiated, a fraction σ_2_ will successfully clear the bacteria at a rate h_2_, corresponding to the treatment duration. We model these patients as transitioning to the susceptible compartment (S) at rate σ_2_h_2_. However, the actual proportion of them that transition to S is different from σ_2_ since multiple outflow processes are being modeled. Individuals in K_1_ remain at risk of developing symptomatic disease. They move to compartment A at a rate *v*K_1_, where *v* is the incubation rate.

#### 1.7 Discharge rates

We estimate the discharge rates from compartments based on hospitalization length data. Non-susceptible, susceptible, and asymptomatic carriers are discharged at rates Ψ_1_, Ψ_2_, and Ψ_3_, respectively, each corresponding to the inverse of the average length of hospital stay. For the infected population, the discharge rate is denoted as Ψ_4_, representing the inverse of the average hospital stay for patients diagnosed with CDI. We describe the movements among compartments by the following differential equations:

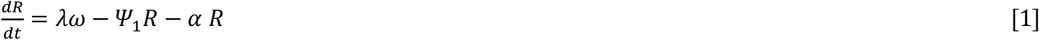

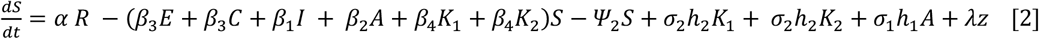

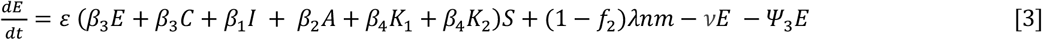

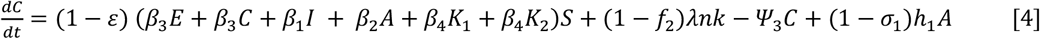

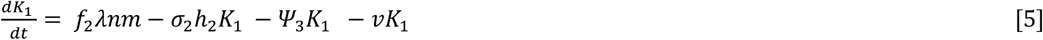

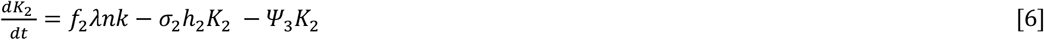

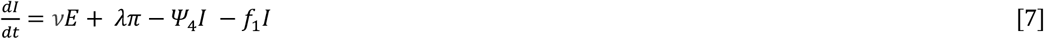

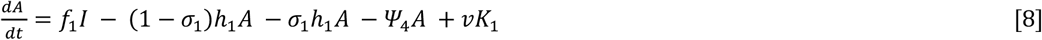

### Model parameters and calibration

Model parameters were obtained from clinical data from the University of California, San Francisco Medical Center, a large tertiary-care, academic hospital, from November 1, 2019, to May 31, 2021. Parameters unavailable in institutional data were derived from previously published studies [8,12,14,17–22] (Table 1).

**Table 1.** Description of input parameters and values used in estimations.

| Symbol | Parameter description (units) | Basal values | Range | Source |
| --- | --- | --- | --- | --- |
| $\delta$ | Transmission rate of non-isolated symptomatic patients (Days<sup>-1</sup>) | 0.00040 | Fixed with data | Fixed with data |
| $\alpha$ | Rate of antibiotic use (Days<sup>-1</sup>) | 0.320 | (0.24-0.38) | Clinical data |
| $\Psi_{1-3}$ | Discharge rate of non-susceptible patients, susceptible patients, and asymptomatic carriers (Days<sup>-1</sup>) | 0.166 | (0.13-0.2) | Clinical data |
| $\Psi_4$ | Discharge rate of symptomatic patients (Days<sup>-1</sup>) | 0.083 | (0.06-0.1) | Clinical data |
| $h_1$ | Bacterial clearance rate due to treatment for infected patients or treatment duration (Days<sup>-1</sup>) | 0.100 | (0.08-0.12) | [12] |
| $\lambda$ | Admission rate (Patients/days) | 76 | - | Clinical data |
| $n$ | Fraction of patients admitted who are asymptomatic carriers (Dless) | 0.08 | (0.06-0.09) | [17,18] |
| $m$ | Fraction of admitted asymptomatic carriers to the E compartment (Dless) | 0.12 | (0.09-0.14) | [18] |
| $k$ | Fraction of asymptomatic carriers admitted to the C compartment (Dless) | 1-m | - | - |
| $\pi$ | Fraction of patients admitted who have symptomatic infections (Dless) | 0.005 | (0.004-0.006)<sup>a</sup> | Clinical data |
| $z$ | Fraction of patients admitted who are not carriers, but are susceptible to colonization (Dless) | 0.22 | (0.17-0.26) | [8] |
| $x$ | Relative transmissibility for asymptomatic carriers (Dless) | 0.700 | (0.56-0.84) | [14] |
| $\varepsilon$ | Fraction of new asymptomatic carriers who may develop symptomatic disease (Dless) | 0.130 | (0.10-0.15) | [18] |
| $v$ | Progression rate to symptomatic disease (Days<sup>-1</sup>) | 0.250 | (0.20-0.30) | [12] |
| $\sigma_1$ | Fraction of cured infected individuals after treatment or treatment effectiveness (Dless) | 0.700 | (0.56-0.84) | [19] |
| $f_1$ | Diagnosis rate of symptomatic patients (Days<sup>-1</sup>) | 0.666 | (0.53-0.80) | [22] |
| $\gamma_1$ | Reduction of transmission due to contact precautions in infected individuals (Dless) | 0.500 | (0.40-0.60) | [20,21] |
| $h_2$ | Bacterial clearance rate due to treatment for asymptomatic patients or treatment duration (Days<sup>-1</sup>) | NA<sup>c</sup> | (0.08-0.12)<sup>b</sup> | [12]<br>Simulated intervention |
| $\sigma_2$ | Fraction of cured asymptomatic carriers after treatment or treatment effectiveness (Dless) | NA<sup>c</sup> | (0.5 - 1.0) | Simulated intervention |
| $\gamma_2$ | Reduction constant of transmission due to contact precautions in asymptomatic carriers (Dless) | NA<sup>c</sup> | (0.5 - 1.0) | Simulated intervention |
| $f_2$ | The proportion of asymptomatic carrier admissions that are immediately diagnosed as carriers | NA<sup>c</sup> | (0 - 1) | Simulated intervention |

We implemented our compartmental model using the deSolve package in R (version 4.3.2). To incorporate uncertainty in parameter values, we conducted 1,000 simulations in which model parameters, except for the transmission rate, were independently sampled from uniform distributions within ±20% of their baseline estimates. The transmission rate or *δ* was optimized in each simulation through numerical calibration by minimizing the absolute difference between the model-predicted cumulative incidence of CDI, quantified as the total number of individuals passing through compartment A, and the observed number of infections over the study period.

^a^Values were derived by determining the proportion of CDI cases in our dataset classified as community-associated CDI. This classification is based on whether or not the time between a CDI diagnosis and the corresponding hospital admission exceeds a time threshold. The baseline time threshold is three days, while the range is one to five days. ^b^In the absence of guidelines, we applied the same range of treatment durations used for CDI patients on colonized individuals. ^c^Parameters related to asymptomatic carrier interventions were not included in the baseline scenario, in which no control measures target this population. Parameter ranges were used in the intervention analysis to evaluate the impact of simulated treatment and/or decolonization strategies (see Methods). Dless: dimensionless

#### Estimation of the Intrinsic Reproduction Number

In our model, individuals with *C. difficile* can either be colonized upon hospital admission or acquire the organism during hospitalization. Both groups can contribute to onward transmission. However, to evaluate the potential for sustained in-hospital transmission independent of importation, we define the intrinsic reproduction number (R_i_) as the average number of secondary cases caused by individuals who became colonized during their hospital stay. For each set of parameters, we calculate R_i_ using the next-generation matrix method [23]. This approach decomposes the system of differential equations into two vector functions:

**ℱ(*x*)**: representing the rate of appearance of new infections in each infectious compartment

**𝒱(*x*)**: representing the transfer rate into and out of each infectious compartment due to all other transitions.

Let *c* denote the vector of infectious compartments. For our model, we define:

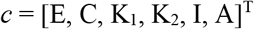

We then compute the Jacobian matrices of *ℱ*(*c*) and *𝒱*(*c*) for the infectious compartments, with entries defined by the partial derivatives:

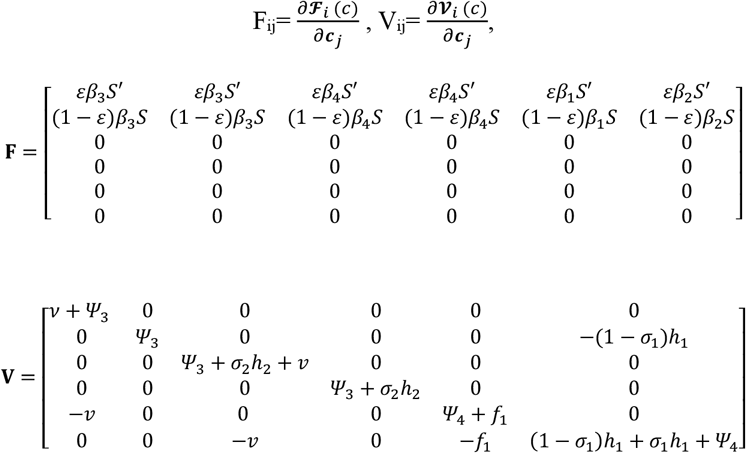

These derivatives were evaluated at the disease-free equilibrium, where all infectious compartments are set to zero and susceptibles are held at their equilibrium value (S = S’):

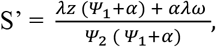

Since this approach focuses on intrinsic self-sustaining transmission to the hospital, rates representing the importation of colonized or infected individuals were not included. The Next Generation Matrix is then defined as:

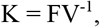

Where the dominant eigenvalue or spectral radius of K corresponds to the intrinsic reproduction number. MATLAB (version R2023a) was used to derive the R_i_ equation.

### Estimation of Colonization Amplification Index

The amplification index, defined as the ratio of hospital-discharged colonized patients to colonized patients at admission under equilibrium conditions, was calculated from the final time point of the simulation. Although the model reached stability early, we conservatively used the last simulated day, when no appreciable changes in the colonization compartments were observed. The total number of colonized individuals in compartments E, C, K_1_, and K_2_ was multiplied by the discharge rate for colonized patients *Ψ*_3_ to estimate the daily number of colonized discharges. This was then divided by the expected daily number of colonized admissions, calculated as λ times *n*:

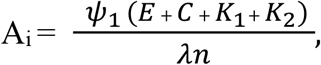

This metric captures the hospital’s contribution to colonization dynamics and complements the R_i_, which reflects transmission intensity. Values of A_i_ greater than one indicate amplification of colonization during hospitalization. Meanwhile, the symptomatic cases are tracked by the overall incidence of CDI.

### Sensitivity Analyses

#### Global Sensitivity Analysis

Partial rank correlation coefficients (PRCC) were used to assess the sensitivity of model outputs to variation in input parameters. As a standard method in global sensitivity analysis, PRCC quantifies monotonic associations between parameters and outcomes, including R_i_ and A_i_, while controlling for the effects of other variables [24]. This method ranks inputs and outputs, and uses multiple regression to estimate the independent contribution of each parameter to the outcome.

#### Impact of Temporal Definitions on Healthcare-Associated Cases

The heuristic classification of CA-CDI directly affects the number of cases classified as HCA-CDI, influencing estimates of in-hospital transmission. The classification of CA-CDI is based on whether the time from hospital admission to symptom onset of CDI is shorter than a specific time threshold.

This threshold is typically set at 3 days for disease reporting standards, but that does not perfectly align with which cases originated in the community [16]. Our model examined how varying this threshold from 1 to 5 days impacted the estimation of R_i_ and A_i_. We used the observed clinical data for each threshold to estimate the proportion of patients admitted with CA-CDI, π, and thus the corresponding number of cases attributed to in-hospital acquisition. The presumptive number of HCA-CDI was then used to estimate the transmission coefficient, *δ*, and the resulting R_i_ and A_i_metrics.

#### Infection Control Interventions Analysis

We extended our baseline transmission model by incorporating additional compartments representing control strategies targeting asymptomatic carriers. Given the uncertainty surrounding poorly defined parameters for colonized individuals, including ɣ_2_, h_2_, and σ_2_, representing intervention effectiveness and treatment rate, we used random sampling from uniform distributions to explore plausible values.

We evaluated how prophylactic treatment and contact precautions for asymptomatic carriers affect CDI incidence and the A_i_ values. Contact precautions aim to limit spread through direct contact [25], while decolonization seeks to eliminate the bacteria, potentially reducing the risk of disease progression and onward transmission. We simulated three scenarios: (1) prophylactic treatment alone, (2) contact precautions alone, and (3) prophylactic treatment combined with contact precautions. For scenario 1, the effectiveness of contact precautions in compartments K_1-2_ was set to zero (ɣ_2_=0), while the treatment completion rate (h_2_) and effectiveness (σ_*2*_) were set to the values provided in Table 2. For scenario 2, where contact precautions were simulated, isolation effectiveness (ɣ_2_) was set to established values, and the treatment completion rate was set to zero (h_2_=0). Lastly, for scenario 3, both interventions were implemented, with the treatment completion rate (h_2_), treatment effectiveness (σ_*2*_), and isolation effectiveness (ɣ_2_) set to their respective values.

## RESULTS

During the study period from November 2019 to June 2021, our healthcare facility recorded a total of 43,320 admissions, with an average of 76 admissions per day. Based on laboratory tests conducted, 4,767 episodes of diarrhea were identified; of these, 659 (13.8%) episodes were confirmed as positive for *C. difficile* toxins via PCR testing. Among the confirmed cases, 10 (1.5%) episodes were considered duplicated due to testing intervals of less than 7 days, and 10 (1.5%) episodes were outpatients, resulting in a final dataset of 639 CDI cases.

According to our records, the overall incidence of CDI was 14.8 cases per 1,000 admissions. Based on the standard case definition specifying that all CO-CDI cases are diagnosed within three days of admission, the HCA-CDI incidence was 10.2 cases per 1,000 admissions, while the CO-CDI incidence was 4.6 cases per 1,000 admissions, with 441 and 198 cases reported, respectively.

Across 1,000 sets of parameters and the standard three-day cutoff for HAI-CDI vs CO-CDI classification, the median colonization amplification index, A_i_, was 1.9 (Q1–Q3: 1.7–2.1), indicating that the number of colonized patients at discharge was approximately 90% higher than at admission. The median intrinsic reproduction number, R_i_, was 0.58 (Q1–Q3 = 0.50–0.65). R_i_ was less than one for all parameter sets, indicating no potential for self-sustaining transmission within the hospital (Fig. 2).

**Fig 2.**
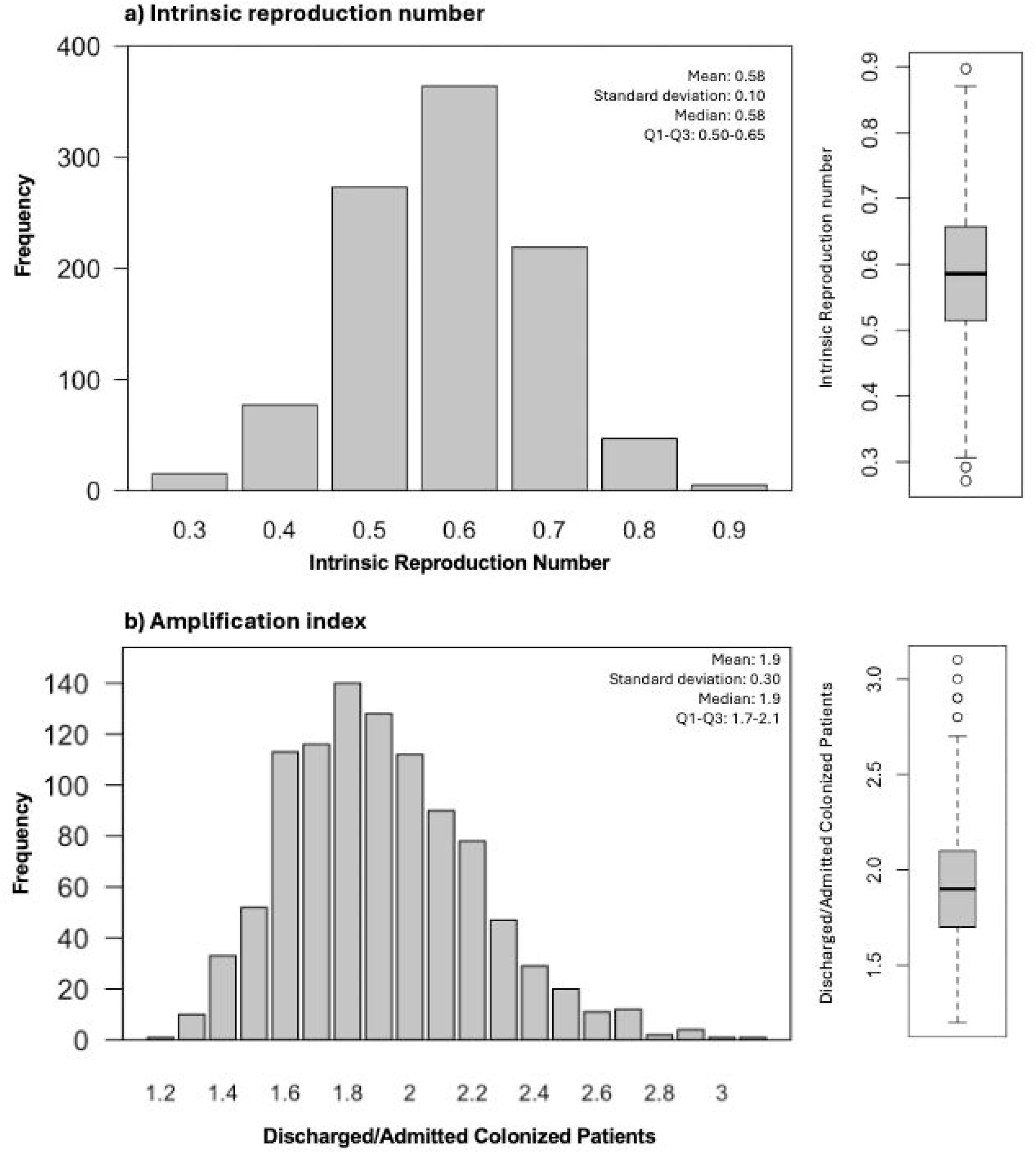
Distribution of the intrinsic reproduction number (R_i_) and the amplification index (A_i_) values. The (a) distribution and box plot of the intrinsic reproduction number, and (b) the colonization amplification index, for 1,000 randomly sampled parameter values. A cutoff of three days was used to distinguish CO-CDI from HCA-CDI.

### Global Sensitivity Analysis

According to the global sensitivity analysis, the transmission rate of symptomatic patients, the relative transmissibility of colonized individuals, and the antibiotic usage rate were the strongest positive predictors of both R_i_ and A_i_ estimates. Conversely, the discharge rates for non-susceptible patients, asymptomatic carriers, and susceptible patients demonstrated the strongest negative correlations. Full PRCC analysis results are presented in S1 and S2 Tables.

### Temporal Definitions on Healthcare-Associated Cases

Our sensitivity analysis explored how the time threshold for distinguishing between CO-CDI and HCA-CDI impacted R_i_ and A_i_ estimates (Fig 3). The median reproduction number increased from 0.53 to 0.68 as the time threshold decreased from five days to one day. Even under the most inclusive definition (1-day threshold), which classified more cases as hospital-acquired, R_i_ remained below one in 99.9% of simulations, suggesting limited potential for sustained transmission. On the other hand, the A_i_ ranged from 1.80 to 2.15 as the time threshold decreased from five days to one day.

**Fig 3.**
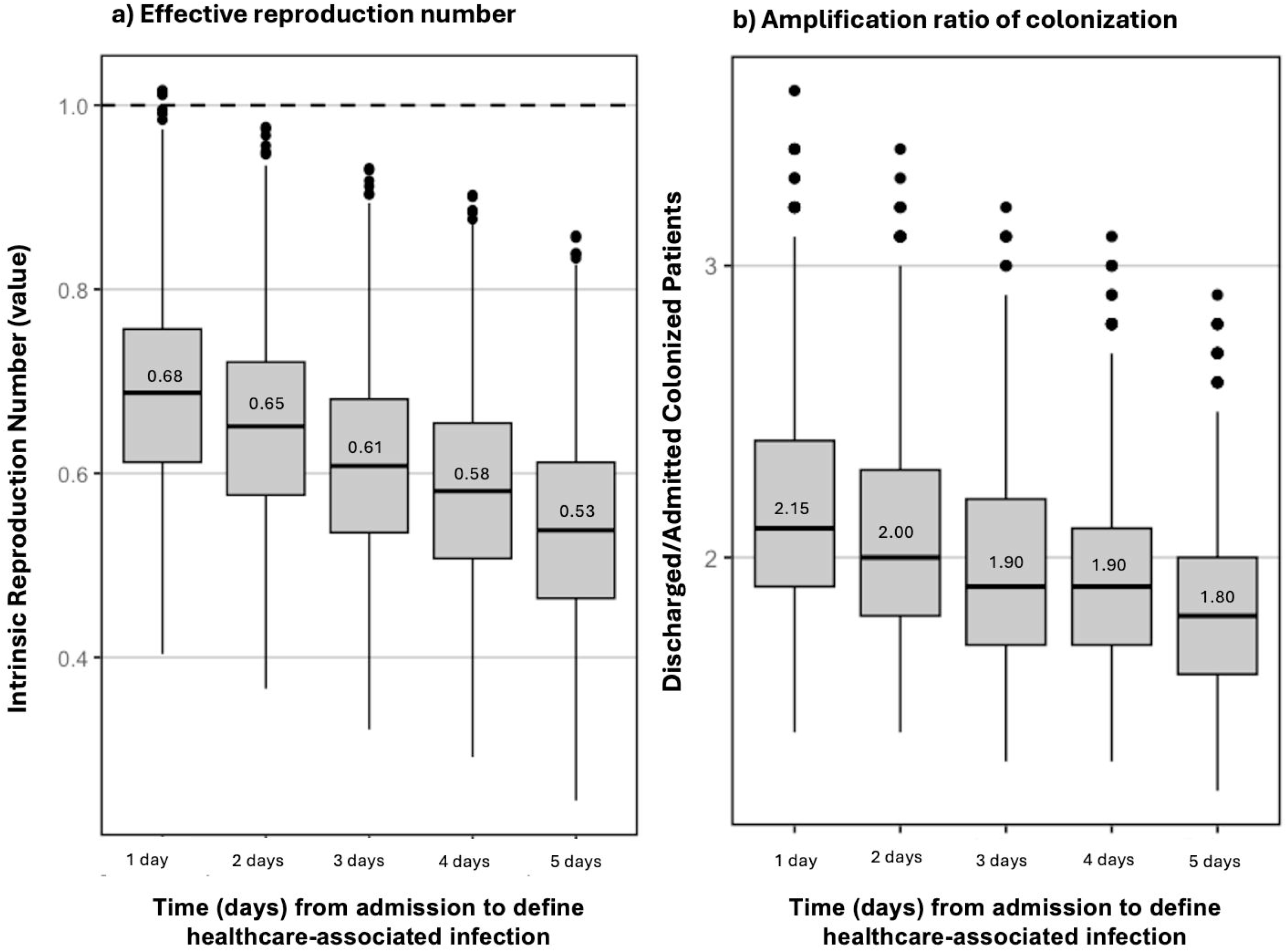
Analysis of Temporal Definitions of Healthcare-Associated Cases. The a) distribution and b) box plot of the baseline number of secondary cases per day in our institution after Monte Carlo simulations with 1,000 random samples.

### Infection Control Interventions Analysis

To assess the impact of contact precautions and prophylactic treatment for asymptomatic carriers, we simulated three intervention strategies: treatment of carriers, isolation of carriers, and both combined (Fig 4). Our results depended on the proportion of patients with asymptomatic carriage who were diagnosed on admission (f_2_) and the effectiveness parameters for isolation (ɣ_2_) and decolonization (σ_2_). As the effectiveness of interventions improves, A_i_ and the overall CDI incidence decrease.

**Fig 4.**
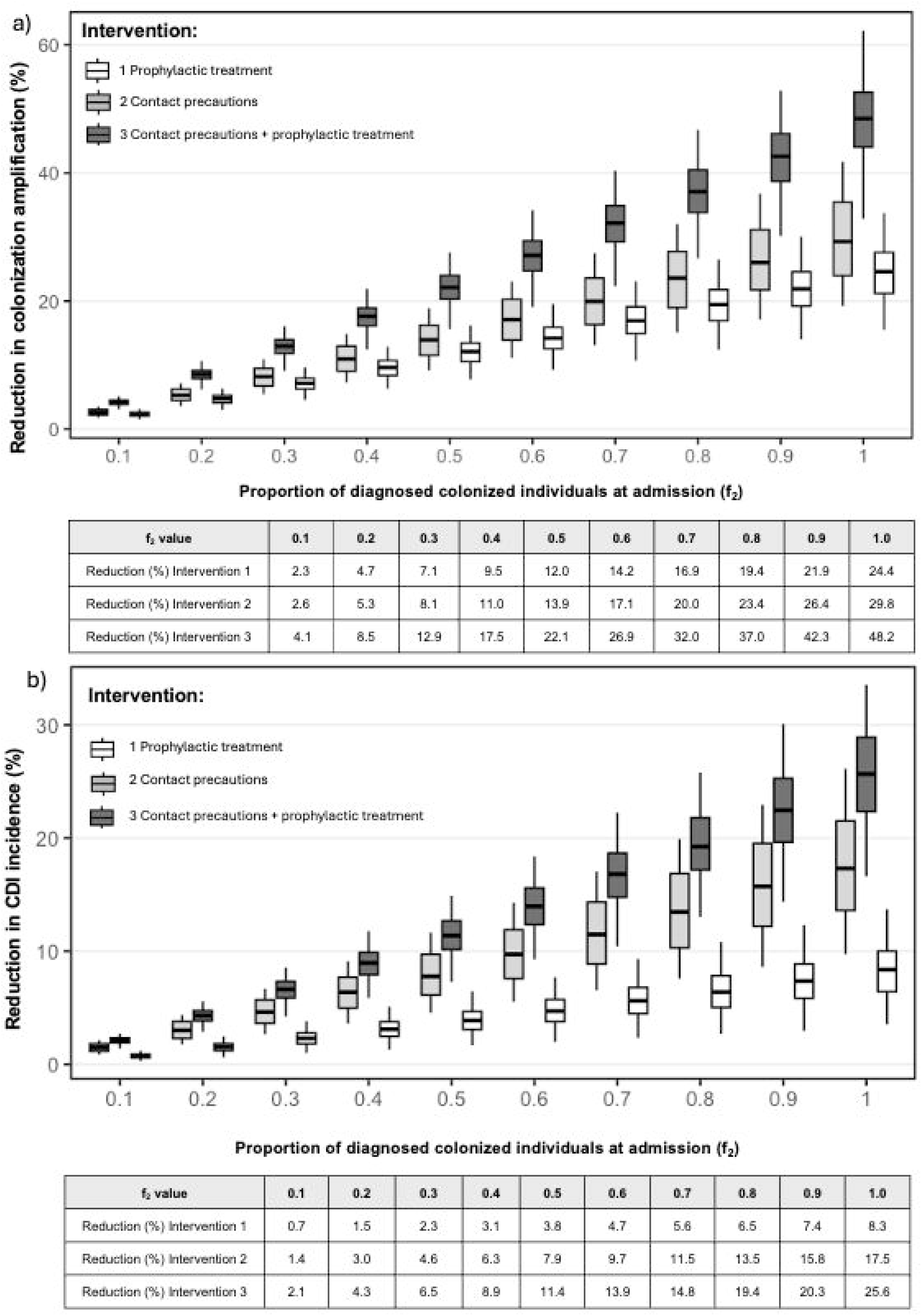
Impact of interventions focused on asymptomatic carriers of *C. difficile*. Changes in the colonization amplification index (top) and CDI incidence (bottom) when applying: 1) prophylactic treatment of carriers at the time of admission, 2) contact precautions for carriers at the time of admission, and 3) prophylactic treatment combined with contact precautions. The boxes in the plots show the 25th to 75th percentile range, with the horizontal line representing the median.

When implemented individually, contact precautions were consistently more effective at reducing A_i_ and CDI incidence than decolonization. Notably, the combined application of both interventions produced the largest effect. For example, isolating and treating 20% of colonized individuals at admission reduced the amplification index by approximately 8.5% and CDI incidence by 4.3%. Combining both interventions yielded an approximately additive reduction in CDI incidence, with the combined effect roughly equal to the sum of their individual effects. However, the decrease in A_i_ was less pronounced, although still approximately additive.

## DISCUSSION

In this study, we developed a compartmental model that incorporated both symptomatic and asymptomatic individuals to quantify *C. difficile* transmission within the hospital setting. Using this framework, we estimated two complementary metrics, the R_i_ number and the A_i_ index, to capture both in-hospital transmission and the hospital’s contribution to community colonization. We further explored the robustness of these estimates through sensitivity analyses of model parameters, assessed the influence of varying case attribution definitions, and evaluated targeted interventions on asymptomatic carriers.

Our findings highlight the bidirectional relationship between hospital and community *C. difficile* burden. Based on our assumptions, the consistently low R_i_ (<1) across scenarios suggests that in-hospital transmission alone does not sustain endemicity. Instead, ongoing importation of cases from the community is necessary to maintain endemicity within healthcare institutions. This highlights the importance of accounting for internal and external dynamics in infection control strategies [26,27].

Meanwhile, hospitals contribute to the broader community burden by discharging colonized individuals, many of whom remain undetected by current screening practices. By introducing the A_i_, our analysis indicates a consistent amplification of colonized cases within the healthcare setting, even in scenarios where R_i_ remains low. These findings align with and complement previous reports documenting increased colonization at discharge, higher risk of CDI among individuals recently exposed to healthcare settings, and studies emphasizing the role of community-hospital interactions [26,28,29]. This hospital-driven amplification may indirectly sustain healthcare-associated transmission if colonized individuals are later readmitted.

Unlike previous studies, our model explicitly quantifies colonization amplification and incorporates interventions targeting asymptomatic carriers. This underscores the need to expand strategies beyond managing symptomatic cases to include interventions targeting asymptomatic carriers at the time of admission and, potentially, at discharge. From a policy perspective, our results suggest that relying solely on R_i_ to evaluate hospital performance may underestimate the broader epidemiological impact of healthcare institutions on *C. difficile* transmission.

Moreover, the sensitivity of R_i_ and A_i_ to heuristic classification of HCA-CDI cases based on the timing of CDI diagnosis underscores how standardized definitions can significantly influence estimates of R_i_. More accurate classification of cases and assessment of healthcare-associated transmission requires consideration of the stochastic nature of disease progression.

Given evidence that asymptomatic carriers may shed *C. difficile* at levels comparable to symptomatic patients, interventions targeting this group, such as contact precautions and antimicrobial prophylaxis, have been proposed as potential strategies to reduce onward transmission [13,14,16]. Our model showed that both contact precautions and prophylactic treatment targeting asymptomatic carriers reduced A_i_ and infection incidence. Contact precautions consistently outperformed prophylaxis, likely due to their immediate effect on interrupting transmission, whereas prophylactic treatment requires time to achieve bacterial clearance, delaying its impact on transmission dynamics. However, unlike prophylaxis, contact precautions do not affect the risk of progression from colonization to symptomatic disease.

Although quasi-experimental studies have supported the effectiveness of contact precautions [30], strategies to prevent the progression from asymptomatic carriage to active infection may be particularly valuable in settings where transmission from asymptomatic carriers is not the primary driver of spread. For example, Miles-Jay et al. showed that in intensive care units (ICUs), only 1% of patients who tested negative for *C. difficile* upon admission acquired the pathogen through cross-transmission, with most cases occurring in previously colonized patients. However, it is essential to note that conditions in ICUs may not reflect those in general hospital wards. ICU patients were typically housed in single rooms and benefited from daily cleaning with sporicidal disinfectants, which likely reduced opportunities for cross-transmission compared to other healthcare settings [31].

There are practical concerns to implementing contact precautions for carriers, including higher costs, potential impacts on patient well-being, and ethical considerations around isolating individuals without symptoms [32]. Although the effectiveness of contact precautions depends on implementation and compliance, current infection control frameworks provide the infrastructure for implementation. Our model can be helpful for cost-effectiveness studies of these interventions by providing estimates of CDI cases and carriers averted.

The relative effectiveness of prophylactic treatment is tempered by studies showing success rates as low as 30–40% in asymptomatic and symptomatic patients [19,33]. Meanwhile, complex microbiome interactions and heterogeneous bacterial clearance rates associated with antibiotic use were not incorporated into our framework and may modify the actual impact of these interventions. Moreover, prophylactic use of antibiotics may carry unintended consequences, including increased risk of secondary infections such as vancomycin-resistant enterococci, thus contributing to the broader challenge of antimicrobial resistance [33,34]. As new decolonization strategies emerge and our understanding of microbiome dynamics improves, our model can be used to ascertain whether targeted or universal decolonization becomes a preferred strategy.

This study has several limitations. First, the biological mechanisms driving colonization and progression to symptomatic infection remain poorly understood, limiting our ability to model these transitions precisely. Second, a compartmental model approach may overlook heterogeneity within populations and individual variations in movement rates. Instead, this approach relies on assumptions of homogeneous mixing and uniform transition rates between compartments. Third, the data used to calibrate the model included a period during the pandemic, which could alter transmission dynamics.

Lastly, the generalizability of our findings should be considered with caution, as specific estimates of R_i_ and A_i_ may vary across institutions depending on local patient demographics, sources of admission, infection control practices, and patterns of antibiotic use. Our model structure captures fundamental and generalizable epidemiological processes underlying *C. difficile* transmission that are applicable to other healthcare settings and can be flexibly adapted with a limited set of parameters. While the aforementioned limitations inspire opportunities for future work, our model provides insight for assessing the transmission dynamics and control of *C. difficile* in healthcare settings.

## Data Availability

All data produced in the present study are available upon reasonable request to the authors

## Acknowledgments

None

## Supporting information

**S1 Table**. Partial rank correlation coefficients for input parameters in the intrinsic reproduction number estimations.

Partial rank correlation coefficients (PRCC) were obtained after Monte Carlo sampling of parameter values. The PRCC provides adjusted correlation values between model parameters and the intrinsic reproduction number. A cutoff of three days was used to distinguish CO-CDI from HCA-CDI.

**S2 Table**. Partial rank correlation coefficients for input parameters in the colonization amplification index estimations.

Partial rank correlation coefficients (PRCC) for the colonization amplification index were obtained after Monte Carlo sampling of parameter values. A cutoff of three days was used to distinguish CO- CDI from HCA-CDI.

