## Supplementary material for "Colonization Amplification despite Limited In-Hospital Transmission: Modeling the Epidemiological Paradox of *C. difficile* and the Impact of Control Strategies in Healthcare Settings": S1 and S2 Tables

**S1 Table. Partial rank correlation coefficients for input parameters in the intrinsic reproduction number estimations.**

| Symbol | Parameter | PRCC | Lower | Upper |
|---|---|---|---|---|
| $\delta$ | Transmission rate of infected patients | 0.921 | 0.897 | 0.945 |
| x | Relative transmissibility for colonized patients | 0.806 | 0.769 | 0.843 |
| $\alpha$ | Rate of antibiotic use | 0.379 | 0.322 | 0.437 |
| z | Fraction of susceptible patients admitted | 0.175 | 0.114 | 0.236 |
| $\Psi_4$ | Discharge rate of symptomatic patients | -0.001 | -0.063 | 0.061 |
| $\gamma_1$ | Reduction constant of transmission due to contact precautions in infected individuals | -0.053 | -0.115 | 0.009 |
| $h_1$ | Bacterial clearance rate due to treatment for infected patients | -0.057 | -0.119 | 0.006 |
| $\sigma_1$ | Fraction of cured infected patients after treatment | -0.067 | -0.129 | -0.005 |
| $f_1$ | Diagnosis rate of infected patients | -0.094 | -0.156 | -0.032 |
| $\nu$ | Progression rate to symptomatic disease | -0.133 | -0.194 | -0.071 |
| $\varepsilon$ | Fraction of asymptomatic carriers who develop symptomatic disease | -0.175 | -0.236 | -0.113 |
| $\Psi_1$ | Discharge rate of non-susceptible patients | -0.356 | -0.414 | -0.298 |
| $\Psi_3$ | Discharge rate of asymptomatic carriers | -0.725 | -0.768 | -0.682 |
| $\Psi_2$ | Discharge rate of susceptible patients | -0.814 | -0.850 | -0.778 |

Partial rank correlation coefficients (PRCC) were obtained after Monte Carlo sampling of parameter values. The PRCC provides adjusted correlation values between model parameters and the intrinsic reproduction number. A cutoff of three days was used to distinguish CO-CDI from HCA-CDI.

**S2 Table. Partial rank correlation coefficients for input parameters in the colonization amplification index estimations.**

| Symbol | Parameter | PRCC | Lower | Upper |
|---|---|---|---|---|
| $\delta$ | Transmission rate of infected patients | 0.912 | 0.886 | 0.937 |
| x | Relative transmissibility for colonized patients | 0.776 | 0.737 | 0.815 |
| $\alpha$ | Rate of antibiotic use | 0.381 | 0.324 | 0.439 |
| z | Fraction of susceptible patients admitted | 0.189 | 0.128 | 0.250 |
| $\nu$ | Progression rate to symptomatic disease | -0.013 | -0.075 | 0.049 |
| $f_1$ | Diagnosis rate of infected patients | -0.029 | -0.091 | 0.033 |
| $\varepsilon$ | Fraction of asymptomatic carriers who develop symptomatic disease | -0.033 | -0.095 | 0.029 |
| $h_1$ | Bacterial clearance rate due to treatment for infected patients | -0.038 | -0.100 | 0.024 |
| $\gamma_1$ | Reduction constant of transmission due to contact precautions in infected individuals | -0.074 | -0.136 | -0.012 |
| $\Psi_4$ | Discharge rate of symptomatic patients | -0.129 | -0.191 | -0.068 |
| $\sigma_1$ | Fraction of cured infected patients after treatment | -0.143 | -0.204 | -0.081 |
| $\Psi_1$ | Discharge rate of non-susceptible patients | -0.365 | -0.422 | -0.307 |
| $\Psi_3$ | Discharge rate of asymptomatic carriers | -0.720 | -0.763 | -0.677 |
| $\Psi_2$ | Discharge rate of susceptible patients | -0.794 | -0.832 | -0.757 |

Partial rank correlation coefficients (PRCC) for the colonization amplification index were obtained after Monte Carlo sampling of parameter values. A cutoff of three days was used to distinguish CO-CDI from HCA-CDI.
